# Quantifying bias from dependent left truncation in survival analyses of real world data

**DOI:** 10.1101/2021.08.02.21261492

**Authors:** Arjun Sondhi, Olivier Humblet, Akshay Swaminathan

**Author notes:** **Corresponding author:** Arjun Sondhi, Flatiron Health Inc., 233 Spring St, New York, NY 10013. **Author contributions:** **Study concept and design:** All **Data collection:** Flatiron Health Inc **Analysis and interpretation:** All **Manuscript writing and review:** All. **Competing interests:**. **Disclosures:** At the time of the study, all authors report employment in Flatiron Health, Inc., an independent subsidiary of Roche. All authors report stock ownership in Roche. **Funding:** This study was sponsored by Flatiron Health, which is an independent subsidiary of the Roche Group.

## Abstract

In real world data (RWD) studies, observed datasets are often subject to left truncation, which can bias estimates of survival parameters. Standard methods can only suitably account for left truncation when survival and entry time are independent. Therefore, in the dependent left truncation setting, it is important to quantify the magnitude and direction of estimator bias to determine whether an analysis provides valid results. We conduct simulation studies of common RWD analytic settings in order to determine when standard analysis provides reliable estimates, and to identify factors that contribute most to estimator bias. We also outline a procedure for conducting a simulation-based sensitivity analysis for an arbitrary dataset subject to dependent left truncation. Our simulation results show that when comparing a truncated real-world arm to a non-truncated arm, we observe the estimated hazard ratio biased upwards, providing conservative inference. The most important data-generating parameter contributing to bias is the proportion of left truncated patients, given any level of dependence between survival and entry time. For specific datasets and analyses that may differ from our example, we recommend applying our sensitivity analysis approach to determine how results would change given varying proportions of truncation.

## 1. Introduction

Recently, the availability of clinical data related to patient health status and/or the delivery of health care has grown substantially. Investigators can tap a variety of routine-care sources (such as claims, registries, electronic health records [EHRs]) to collect these data observationally; therefore, their architecture is not pre-specified as it would be in clinical trials. These types of data have been termed real world data (RWD), and their handling and analysis requires particular analytic considerations. In many electronic health record (EHR) databases, patients are only observed if they satisfy certain entry criteria, such as having a specific number of clinic visits, or undergoing a biomarker testing procedure.^1,2^ This also occurs in prospective cohort studies where patients are enrolled after an initiating event of interest.^3^ In time-to-event analyses, if a patient enters the study or database after the start of their follow-up or index time, they are said to have *delayed entry*. With this sampling process, we do not observe any patients who experienced a disqualifying event (such as death) before satisfying the entry criteria, which is referred to as *left truncation*.^4^ Failure to account for left truncation by including patients in a survival analysis at their index time and accruing person-time prior to satisfying the entry criteria results in a selection bias, since patients observed in the database had to at least live long enough to qualify for entry.^5,6^

When dealing with left truncated data, standard methods for estimating the marginal survival distribution are instead estimating survival conditional on surviving up to entry time. The most common approach for recovering the marginal distribution is *risk set adjustment*, where patients are only considered to be at risk for the endpoint of interest once they have satisfied the entry criteria. In contrast, standard methods treat all patients as at risk from the start of their follow-up or index time until their observed event or censoring time, resulting in bias. Risk set adjustment is easy to implement with standard software, and can be used with both Kaplan-Meier survival estimators and Cox proportional hazards regression models.^7^ However, it relies on the assumption that the time to the endpoint T is independent of the time to entry E, given that T > E. In other words, patients who entered later should have the same hazard of death as those who entered earlier. This *independent left truncation* assumption is testable. The simplest method is the coefficient test of a risk set adjusted Cox model with entry time as the sole covariate.^8^ There are also tests based on conditional Kendall’s tau that may be more appropriate when the Cox model assumption does not hold.^9 10^

In practice, however, the assumption of independence is not always satisfied. For example, in a study of mutation-positive patients with cancer identified through biomarker testing, it is possible that testing could be triggered by worsening of disease and exhaustion of standard therapies, or that patients tested earlier have better outcomes due to timely receipt of targeted therapies. If there is dependency between survival and entry time, then risk set adjustment is no longer guaranteed to unbiasedly estimate the marginal survival distribution. However, the magnitude and direction of this bias is generally unexplored in the literature. While there have been some proposed methodological solutions for dependent left truncation, these make strong parametric assumptions that are untestable from observed data, and generally do not extend to multivariate regression modeling.^11^

In this work, we design and implement simulation studies of analyses involving survival data subject to dependent left truncation, based on studies with real world clinico-genomic data, where patients are only observed upon receiving a genomic test, and survival is typically measured from the start of a certain treatment. Given the lack of commonly-accepted methods for dependent left truncation, we quantify the inferential bias that occurs when fitting risk set adjusted Cox regression and Kaplan-Meier estimators. We characterize general trends in the bias, with the goal of determining when such analyses may still provide useful results. In addition, we provide a procedure for conducting sensitivity analyses given an arbitrary dataset subject to dependent left truncation. This can be used by analysts to determine how bias in results would change when varying non-estimable parameters.

## 2. Methods

In this section, we apply a simulation approach to study the bias induced from dependent left truncation when applying standard and risk set adjusted methods. We first describe how to design a simulation study informed by a particular dataset, where most parameters can be estimated and others can be varied as a sensitivity analysis. We then apply this framework to conduct a simulation study; two additional analytic settings can be found in the Supplement.

### 2.1 Simulation-based sensitivity analysis

The goal of our method is to assess the expected bias due to dependent left truncation when estimating parameters of interest. We conduct the following steps to create a data-generating process that is congenial with the observed dataset:

- Assume generative models for survival, entry, and censoring
- Estimate parameters from observed dataset
- Vary parameters that are not estimable
- Perform simulation study

Most parameters in the simulation data-generating process can be estimated in order to match the dataset of interest. However, we vary the distributions of truncation and censoring across a range of parameters, since these are not estimable from dependently truncated data.

For illustrative purposes, we assume a survival analysis comparing two treatments, with both data arms subject to dependent left truncation. The estimand of interest is the marginal hazard ratio comparing the two treatments. For simplicity, we further assume that treatment is unconfounded, although confounding variables can easily be added to the generative models for treatment arm assignment and survival time.

#### Assume generative models

The first step is to assume the form of the generative models that created the dataset. We describe a simple survival model that matches what we use in our simulation study described in the next section, although more complicated forms can also be used. Specifically, we assume

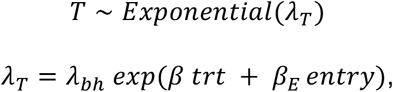

where *λ*_*T*_ is the hazard function for *T*the time to death, composed of the constant baseline hazard *λ*_*bh*_ with parameters *β*and *β*_*E*_ describing the conditional survival-treatment and survival-entry associations. We then assume the following models for treatment and entry times:

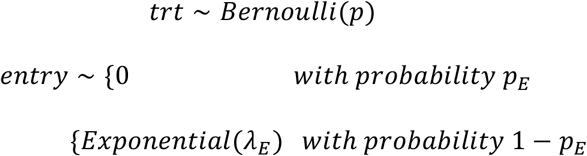

where*p, λ*_*E*_and*p*_*E*_are constant. Finally, we assume a model for censoring:

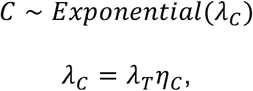

where *λ*_*C*_ is the hazard function for *C*the time to censoring, and the parameter *η*_*C*_results in a censoring probability of *η*_*C*_/(1 + *η*_*C*_) in the non-truncated dataset. Then, the time-to-event is*Y* = *min*(*T, C*). Truncation is applied by filtering out observations where *Y* < *entry*.

#### Estimate parameters from dataset

Next, we estimate as many parameters for this generative model as we are able to do from our observed truncated dataset. Under our proportional hazards model assumption, the conditional model of survival time as a function of treatment arm and entry time is estimable; that is, estimates of *λ*_*bh*_, *β*, and *β*_*E*_ can be obtained by fitting a risk set adjusted Cox model with treatment and entry time as regressors. Additionally, the probability of treatment *p* is readily estimable from the observed dataset, under our generative model for treatment, which is independent of entry time. Finally, the probability of pre-index entry to the cohort conditional on not being truncated, *P*(*entry* = 0 | *Y* > *entry*), is also estimable from the observed dataset. By conditional probability, we have that *p*_*E*_ = *P*(*entry* = 0 | *Y* > *entry*)*P*(*Y* > *entry*), which allows us to recover an estimate for *p*_*E*_once we set *P*(*Y* > *entry*)as described below.

The parameters *η*_*C*_and *λ*_*E*_, which characterize the censoring and entry time distributions, are not estimable from observed data, and therefore must be varied in simulation. We consider these as sensitivity parameters.

#### Vary parameters that are not estimable

Recall that the parameter of interest is the marginal hazard ratio comparing the two treatments. This would be estimated by fitting a standard Cox model on the non-truncated dataset, with treatment indicator as the sole regressor. However, because we only observe the truncated dataset, this parameter is not estimable under dependent left truncation. Under the data generating process, the marginal hazard ratio is a function of the entry time distribution.

Therefore, as we vary *λ*_*E*_, we are obtaining different true marginal hazard ratio parameters. Varying *η*_*C*_would affect the estimate of the marginal hazard ratio based on the non-truncated dataset.

We can select values for *λ*_*E*_by choosing a parameter value that is more interpretable: the probability of a patient being truncated in the dataset. Each value of *λ*_*E*_corresponds to a distinct entry time distribution, and therefore a distinct *P*(*Y* < *entry*)or truncation probability, assuming a fixed distribution of*Y*. We can then treat *P*(*Y* < *entry*)as the simulation parameter of interest, and select *λ*_*E*_such that a given truncation probability is achieved. Because of the dependency of *Y*on *entry*, this probability is difficult to obtain in closed-form. However, it can be estimated by simulating many datasets with the same *λ*_*E*_and computing the mean observed truncation proportion. Given a function that implements this procedure, a root-finding algorithm can then be applied in order to solve for the corresponding *λ*_*E*_for a certain *P*(*Y* < *entry*). Larger values of the sensitivity parameters *η*_*C*_and *P*(*Y* < *entry*) respectively correspond to greater prevalence of censoring and truncation in the population. Plausible ranges for these should be determined based on domain knowledge or external data. Then, for each parameter configuration, a simulation study can be performed to estimate the bias due to dependent left truncation.

#### Perform simulation study

For a particular data-generating process, we repeat the following steps over a large number of iterations:

- Generate non-truncated dataset
- Compute true parameter of interest
- Apply truncation and obtain truncated dataset
- Fit models to estimate parameter of interest
- Compute error metrics

#### Extensions

Though we have illustrated simulating a simple data-generating process here, it is straightforward to extend to more complex settings. If we suspected that there were differing levels of truncation between the arms, we could define separate entry time distributions, and vary both the corresponding truncation probabilities. Similarly, the dependence between survival and entry time can be allowed to differ between arms by adding an interaction term between treatment and entry time to the hazard function for *T*. The entry time or other covariate associations with survival could also be modeled more flexibly, such as with spline terms.

Additionally, while we assumed unconfounded treatment in the simulation data-generating process described above, this method can be extended to handle confounded treatment assignment. A measured confounder (or vector of confounders) *X* can be added to both the hazard function for *T*, and the model for probability of treatment. The relevant conditional regression parameters are estimable from the observed truncated data, and can be used for observational causal inference procedures. The subsequent simulation can proceed by incorporating e.g. inverse propensity score weighting into both the non-truncated and truncated analysis.

More flexible distributions can also be specified for the random variables in this framework. Although we described survival times drawn from an exponential distribution, this is not strictly necessary. To unbiasedly estimate the survival distribution conditional on entry time, we only require the proportional hazards assumption to hold. Then, given the estimated regression component of the hazard function, more flexible modeling can be used for the baseline hazard. For example, one could fit more complex parametric distributions, or a spline model, as implemented in the *simsurv* R package. It is then straightforward to simulate survival times. However, it is important to note that having entry time drawn from a single parameter distribution allows for the 1:1 correspondence between the parameter and the truncation probability. A distribution with more parameters would lead to more sensitivity parameters that may be less interpretable.

### 2.2 Simulation study of dependent left truncation

In this section, we describe a simulation study conducted to characterize the bias induced through dependent left truncation in a real world control arm that is compared to a non-truncated treatment arm. This comparison of a non-truncated treatment arm to a real world control arm (rwCA) demonstrates a potential use for real world EHR-derived data^12,13^ where a treatment arm from e.g. a clinical trial is not subject to any delayed entry or left truncation, while the real world control arm used for comparison is. The estimand of interest is the marginal hazard ratio of death comparing the two treatments. We also considered two additional designs corresponding to common survival analyses performed with real world data subject to left truncation, which we have detailed in the Supplement.

**Table 1.** Summary of simulation designs implemented, with truncation mechanisms and parameters of interest specified.

|  | Simulation design |  |  |
| --- | --- | --- | --- |
|  | NT arm + rwCA | Two RW arms | Single rwA |
| Treatment arm | No LT | LT | LT |
| **Control arm** | LT | LT |  |
| **Estimand** | Hazard ratio | Hazard ratio | Median survival time |

We describe the parameters used below. In general, these were set to realistic real-world values based on corresponding parameters observed in the nationwide (US-based) de-identified Flatiron Health-Foundation Medicine Clinico-Genomic Database. The de-identified data originated from approximately 280 US cancer clinics (∼800 sites of care). Retrospective longitudinal clinical data were derived from electronic health record (EHR) data, comprising patient-level structured and unstructured data, curated via technology-enabled abstraction, and were linked to genomic data derived from FMI comprehensive genomic profiling (CGP) tests in the FH-FMI CGDB by de-identified, deterministic matching.^14^ Genomic alterations were identified via comprehensive genomic profiling (CGP) of >300 cancer-related genes on FMI’s next-generation sequencing (NGS) based FoundationOne® panel.^15,16^ To date, over 400,000 samples have been sequenced from patients across the US.

As in Section 2.1, we begin with the following model for survival time:

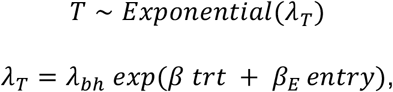

where *λ*_*T*_ is the hazard function for *T*the time to death, composed of the constant baseline hazard *λ*_*bh*_with parameters *β*and *β*_*E*_ describing the conditional survival-treatment and survival-entry associations. The parameters were then set as follows:

- We set *λ*_*bh*_ to be 1/12, which corresponds to an average survival time of 12 months for patients on the *trt* = 0 arm who entered the cohort at their index time.
- The parameter *β*was set to *log*(0.8), which implies that patients on the *trt* = 1arm have 80% of the hazard of death compared to patients on the *trt* = 0arm who entered the cohort at the same time.
- *β*_*E*_ is the parameter describing the association between survival and entry time, i.e. the dependency of the left truncation mechanism. We varied this parameter among *log*(1, 1.01, 1.03, 1.05, 1.10).

We then use the following models for treatment and entry times, where patients with *trt* = 1are not subject to any delayed entry, and therefore cannot be left truncated:

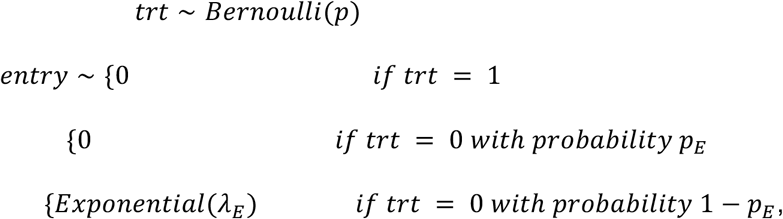

and set the parameters as:

- *p* is fixed at 0.5
- *p*_*E*_ is fixed at 0.2
- *λ*_*E*_ is varied among values that resulted in left truncation probabilities *P*(*Y* > *entry* | *trt* = 0) of 0.1, 0.2, …, 0.7, given all the other parameters set

Finally, we set the censoring model as

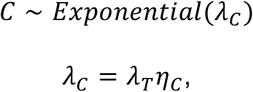

where *λ*_*C*_ is the hazard function for *C*the time to censoring, and the parameter *η*_*C*_results in a censoring probability of *η*_*C*_/(1 + *η*_*C*_) in the complete dataset (before truncation is applied). We fixed *η*_*C*_ = 1.

## 3. Results

In this section, we report the results of our simulation study conducted for one analytic setting; results from the additional simulation studies can be found in the Supplement. These reflect the parameter settings described in Section 2. Varying other parameters in the data-generating process (such as *p*_*E*_, *p, λ*_*bh*_, and *β*) did not meaningfully affect results.

In Figure 1, we display the coverage probability results for this setting with the same parameters as used previously; here, the truncation probability only applies to the real world control arm. We see that 95% confidence intervals for the risk set adjusted Cox hazard ratio provide valid coverage up to a left truncation prevalence of 0.4 at the highest strength of dependency. This implies that differential left truncation between treatment arms results in a more difficult parameter to estimate. The naive estimator performs much worse here, notably also given independent left truncation.

**Figure 1.**
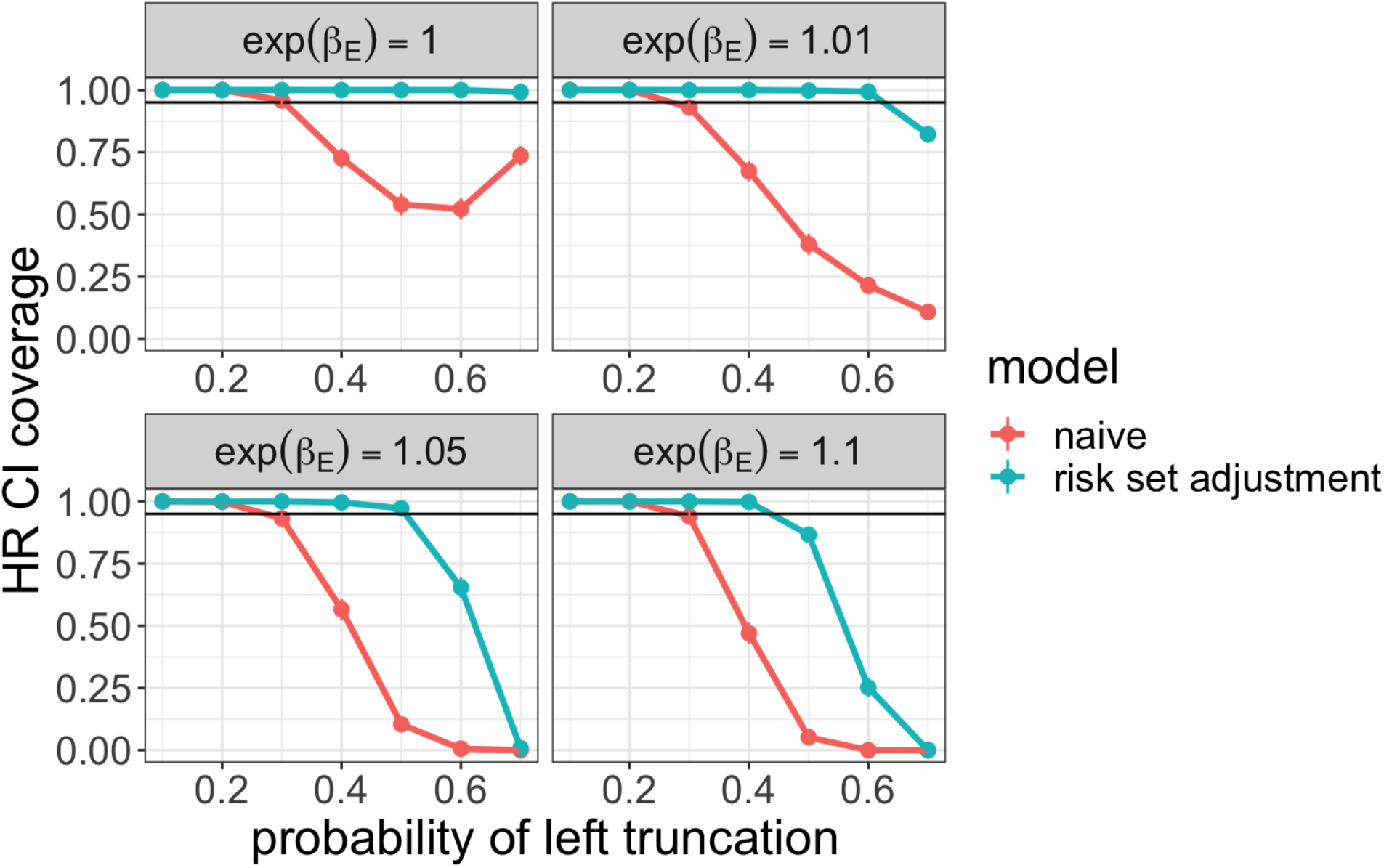
Coverage probability of 95% confidence intervals for treatment hazard ratio comparing a non-truncated arm to a real world arm across simulation settings.

Next, we display the relative bias results for the simulation parameters in Figure 2. As in the first setting, we see that risk set adjustment is unbiased with independent left truncation, and bias increases with truncation and dependency. An interesting result here is that the bias is uniformly upwards, meaning that the estimated hazard ratio is always larger than the true hazard ratio. This phenomenon makes sense in the presence of dependent left truncation. Intuitively, we expect observed patient entries would be earlier compared to the non-truncated entry time distribution. Then, given our data generating process, where later entry time is negatively associated with survival, these patients are also more likely to have a lower hazard of death. Therefore, the estimated hazard ratio comparing the non-truncated treatment arm to the dependently truncated arm will be artificially inflated.

**Figure 2.**
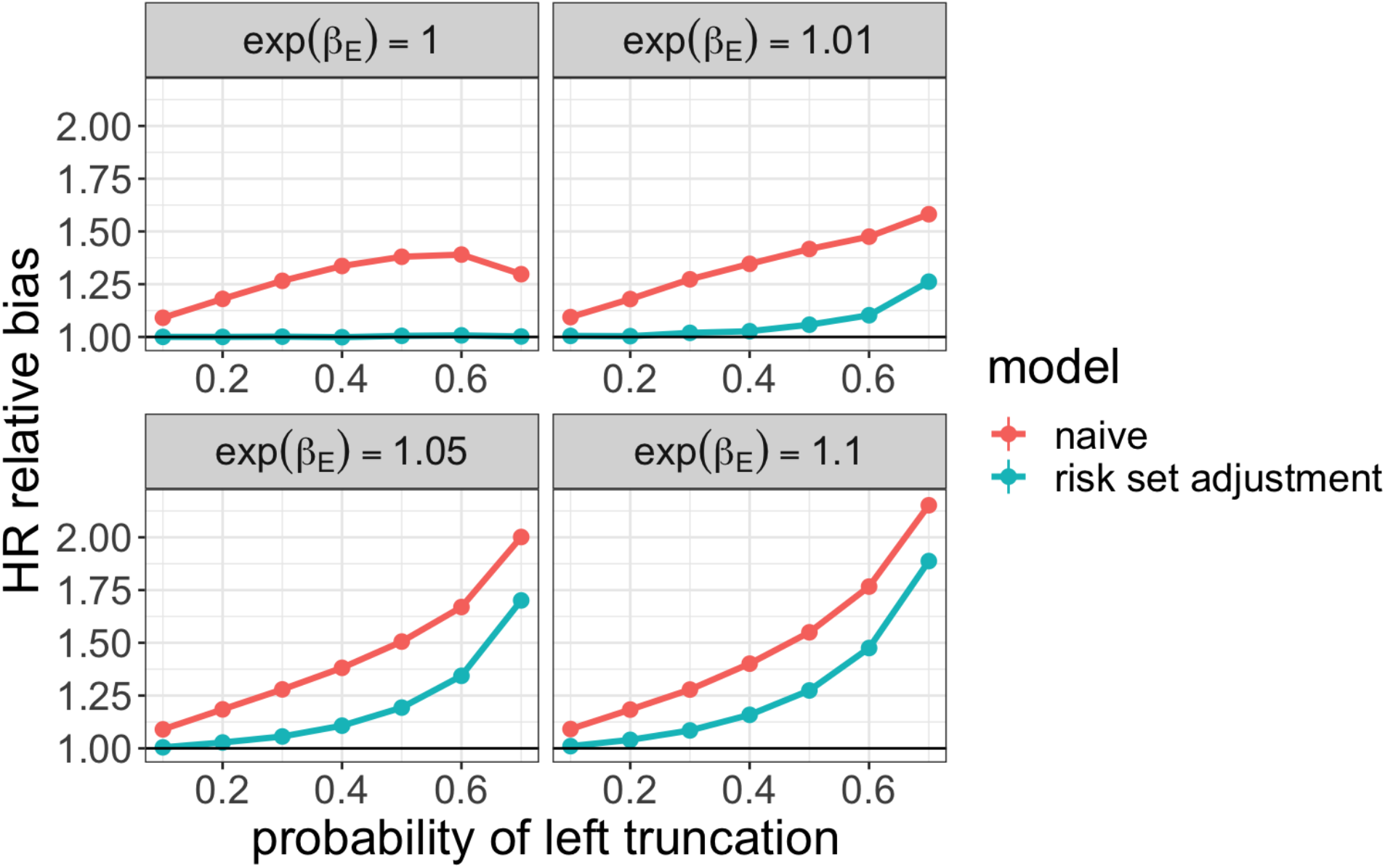
Relative bias for estimated treatment hazard ratio comparing a non-truncated arm to a real world arm across simulation settings.

Recall that the true hazard ratio is less than 1, implying a protective effect of the treatment over the control. In many applications, we would expect that this is the substantive hypothesis being tested; therefore, since the estimate is biased towards the null hypothesis, inference will be conservative.

## 4. Discussion

Through our simulation studies, we measured the impact of dependent left truncation on estimator bias. We reported results with varying probabilities of left truncation (i.e. the proportion of patients who were not observed in the sample due to being truncated) and dependence between survival and entry time; varying other data-generating parameters did not meaningfully affect estimator bias. We found that results varied greatly between analytical settings and estimands, with relative parameters (hazard ratios) being subject to less bias than absolute parameters (median survival time, as shown in the Supplement). At the highest level of dependency assessed (entry time HR of survival = 1.1), confidence intervals for median survival time only had valid coverage up to 0.3 probability of truncation. Hazard ratio confidence intervals provided valid coverage more often, particularly when the level of truncation and survival-entry time dependency in both arms was the same (see Supplement). Even with valid coverage, however, bias can be substantial in point estimates subject to dependent left truncation.

When patients in a non-truncated treatment arm given a protective treatment are compared to a dependently truncated cohort treated in a real world setting, the resulting bias in the hazard ratio is directionally upwards, or towards the null. Specifically, this occurs when later entry time is associated with increased hazard of death, which is empirically true in the clinico-genomic setting of the database used for this report, and is expected, given the survivorship bias (i.e. artificially lower hazard in the truncated cohort) that cannot be corrected via risk set adjustment. The resulting inference is then conservative since the treatment effect is more likely to be declared non-significant. Therefore, if a non-truncated treatment arm shows a significantly lower hazard of death compared to the real world arm, a decision to advance the treatment development can be made with confidence. In drug development, conservative inference is usually preferable to the bias being directionally away from the null, which would lead to a greater likelihood of false positive results. We therefore conclude that there exist analyses where bias due to dependent left truncation may be acceptable in direction or in magnitude. Our simulation approach shows general trends in estimator performance given dependent left truncation; however, the settings will not apply to all possible data analyses. Therefore, we also generalized this simulation study into a sensitivity analysis procedure that can be applied to any arbitrary dataset subject to dependent left truncation. Under the Cox proportional hazards model assumption, many of the corresponding generative model parameters are estimable from data; the parameters determining the censoring and truncation probabilities are generally not. These can instead be varied across a reasonable grid determined by domain knowledge or comparisons to external datasets that are not truncated. Then, for each parameter configuration, the simulation procedure can estimate the level of bias expected to then inform how accurate the initial analysis on the observed dataset is. This method applies to a variety of common estimands, including hazard ratios (that may be conditional on baseline covariates, or marginal over the target population) or median survival time. We recommend that this approach be used by analysts to better quantify uncertainty around results when dependent left truncation is present.

A limitation of our work is that accurate bias quantification requires knowledge of the population truncation proportion, which is not estimable from a truncated dataset. We treat this as a varying sensitivity parameter, which results in a range of bias estimates. However, this range may be too wide to provide useful information. In practice, we suggest that plausible estimates of the truncation proportion be determined through domain knowledge or external data that is not subject to truncation. Our method also does not provide guidance as to what level of bias is considered low enough in order to draw conclusions from an analysis subject to dependent left truncation; this would vary among different applications. An expected relative bias of 10% in the treatment hazard ratio may not affect decision making in a phase I single arm trial, but could be of greater concern for a final regulatory decision.

This study is the first to our knowledge, to quantify the bias resulting from applying standard survival analysis techniques (i.e. Cox proportional hazards models and Kaplan-Meier estimators) with risk set adjustment to left truncated data. Although more complex models have been proposed to account for dependent left truncation, they require unverifiable parametric assumptions, are not commonly used in practice, or do not extend to regression modeling.^11,17^ Another common strategy for handling left truncated data is to restrict the sample to only include subjects who satisfied the conditions for cohort entry before their index time. This eliminates delayed entry (and therefore left truncation) by design, and resulting survival analyses are no longer subject to bias. However, this changes the parameter being estimated; rather than estimating marginal hazard ratios or survival distributions, we are now estimating parameters conditional on entry before index. This results in inference on a population which may not be of scientific interest. In summary, in light of the lack of available methods for mitigating bias from left truncation, it is of interest to determine how reliable standard methods are for estimating marginal parameters under dependent left truncation.

## Data Availability

The data that support the findings of this study have been originated by Flatiron Health, Inc. These de-identified data may be made available upon request, and are subject to a license agreement with Flatiron Health; interested researchers should contact to determine licensing terms

## Appendix/Supplementary

### Two real world treatment arms

As in Section 2.1, we begin with the following model for survival time:

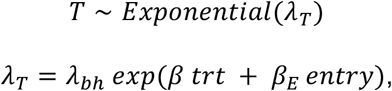

where *λ*_*T*_ is the hazard function for *T* the time to death, composed of the constant baseline hazard *λ*_*bh*_ with parameters *β*and *β*_*E*_ describing the conditional survival-treatment and survival-entry associations. The parameters were then set as follows:

We then use the following models for treatment and entry times

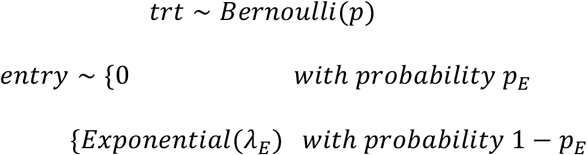

and set the parameters as:

- *p* is fixed at 0.5
- *p*_*E*_ is fixed at 0.2
- *λ*_*E*_ is varied among values that resulted in left truncation probabilities *P*(*Y* > *entry*) of 0.1, 0.2, …, 0.7, given all the other parameters set

Finally, we set the censoring model as

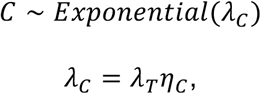

where *λ*_*C*_ is the hazard function for *C*the time to censoring, and the parameter *η*_*C*_results in a censoring probability of *η*_*C*_/(1 + *η*_*C*_) in the non-truncated dataset. We fixed *η*_*C*_ = 1.

Figure S1 displays the coverage probability for the Cox model estimated treatment hazard ratio, both with and without risk set adjustment. We vary the left truncation dependency parameter from 1 (i.e. independent left truncation) to 1.1, which corresponds to a 10% increase in hazard of death for each month later in entry time after follow-up. For each of these, we display the coverage probability across simulations for each probability of left truncation.

In both independent and dependent left truncation settings, valid confidence interval coverage (i.e. at least 95%) is obtained throughout all levels of truncation. Interestingly, this holds for both the risk set adjusted model and the naive model that does not account for delayed entry. This may be because both real world arms were subject to an equal amount of truncation, resulting in low bias for hazard ratio estimation.

**Figure S1.**
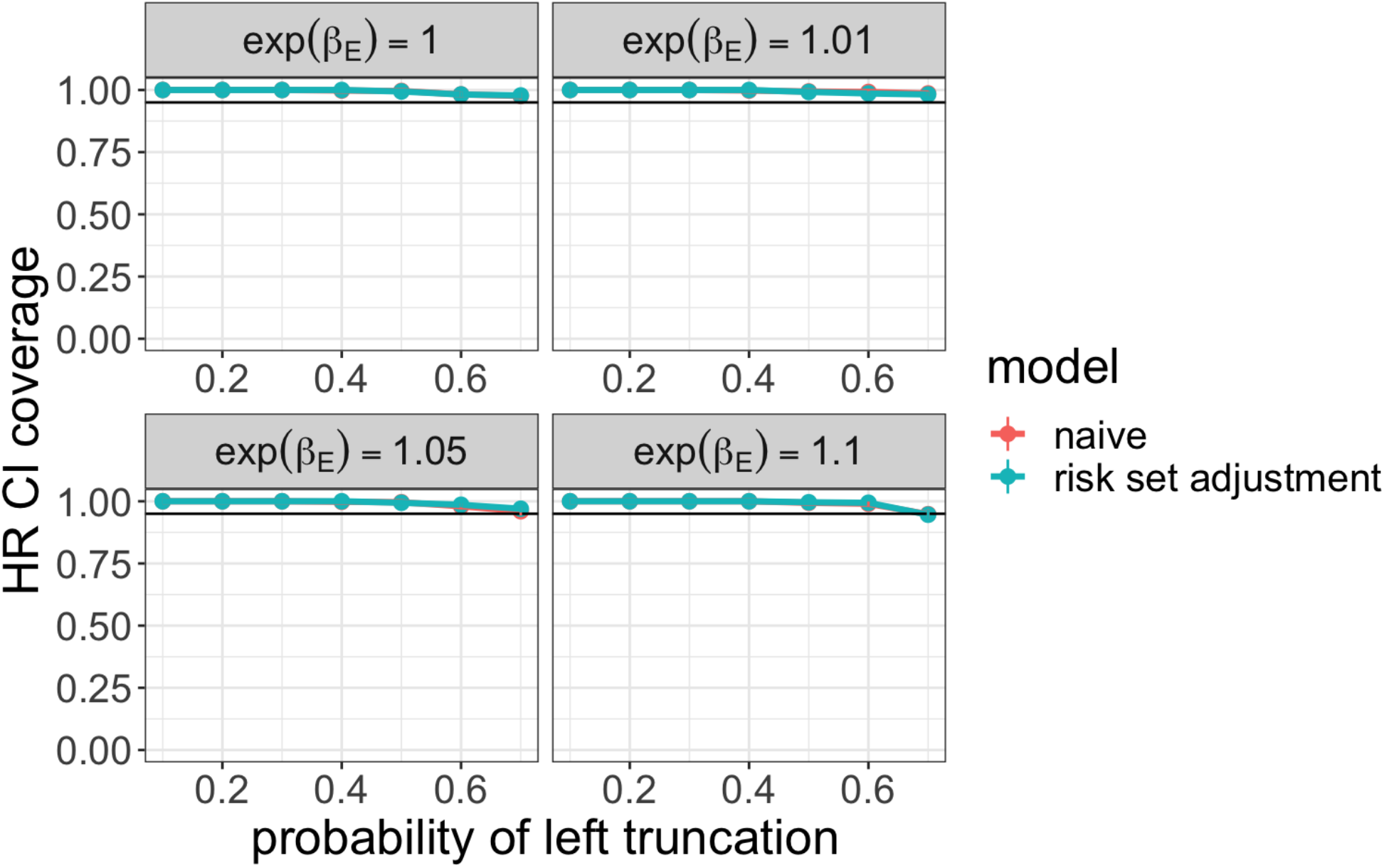
Coverage probability of 95% confidence intervals for treatment hazard ratio comparing two real world treatment arms across simulation settings.

In Figure S2, we show the relative bias of the estimated hazard ratios across the same simulation parameters. In spite of the valid coverage, we observe some bias in the point estimates, which reach up to a 9% under-estimate of the true treatment hazard ratio. With dependency and truncation increasing, we see increased bias in both estimators, though the risk set adjusted model uniformly outperforms the naive model, as expected. This indicates that even with valid coverage, caution is warranted in interpreting the point estimate of the hazard ratio. It is also important to note that the bias is consistently downwards (away from the null), which leads to anti-conservative inference for the parameter.

**Figure S2.**
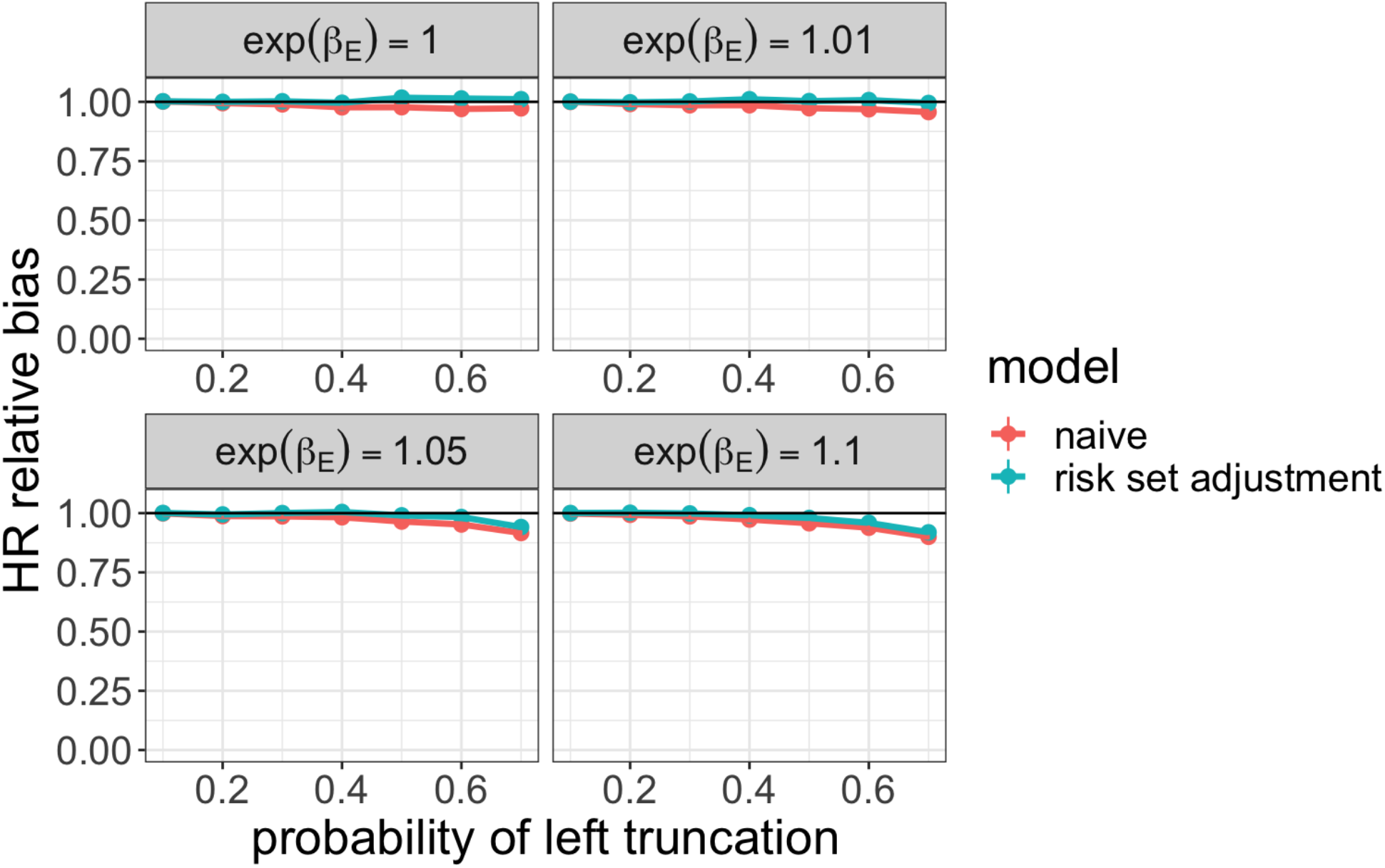
Relative bias for estimated treatment hazard ratio comparing two real world treatment arms across simulation settings.

In this simulation, the truncation dependency and entry time distribution are the same in both arms, and the survival distribution only differs due to treatment. Our results should therefore only be interpreted in this context. In real world data, we may expect that populations may receive different treatments, and also have different mechanisms of cohort entry. To encode these characteristics, the generative model can be modified, as described in Section 2.1.

### Single real world arm

The model for survival time is slightly different for this setting with only a single treatment:

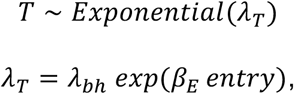

The parameters were then set the same as in the previous settings, as is the censoring model. There is no treatment assignment model for this setting, and the entry time model is

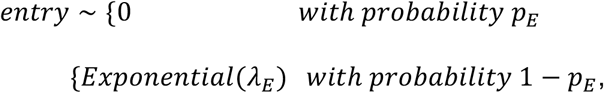

with the following parameters:

- *p*_*E*_ is fixed at 0.2
- *λ*_*E*_ is varied among values that resulted in probabilities of left truncation across the single arm of 0.1, 0.2, …, 0.7, given all the other parameters set

For each unique parameter configuration, we conduct 500 simulation iterations. For each iteration, we fix the truncated dataset sample size at 300 (corresponding to 150 patients observed in each arm for the first two settings), and generate a larger non-truncated dataset according to the truncation probability parameter. The observed time-to-event is*Y* = *min*(*T, C*). Truncation is applied by filtering out observations where *Y* < *entry*. The “true” parameter of interest is defined to be the hazard ratio or median survival time estimate computed on the non-truncated dataset. We compare this to the estimate based on the truncated dataset and report two metrics across simulation iterations: (1) average relative bias, defined as the average ratio of the estimated parameter to the true parameter and (2) coverage probability, the proportion of iterations where the 95% confidence interval for the estimated parameter covers the true parameter.

Finally, we evaluate both naive and risk set adjusted Kaplan-Meier estimation of median survival time given data from a single treatment arm subject to left truncation. We display the coverage results in Figure S3.

Here, we observe a much more dramatic drop-off in coverage, both with increasing dependency and with increasing truncation prevalence. Here, 95% confidence intervals for the risk set adjusted Kaplan-Meier estimate of median survival time provide valid coverage only up to a prevalence of 0.3. When compared to the previous settings, this shows that estimating an absolute quantity is a much more difficult task in the presence of dependent left truncation.

**Figure S3.**
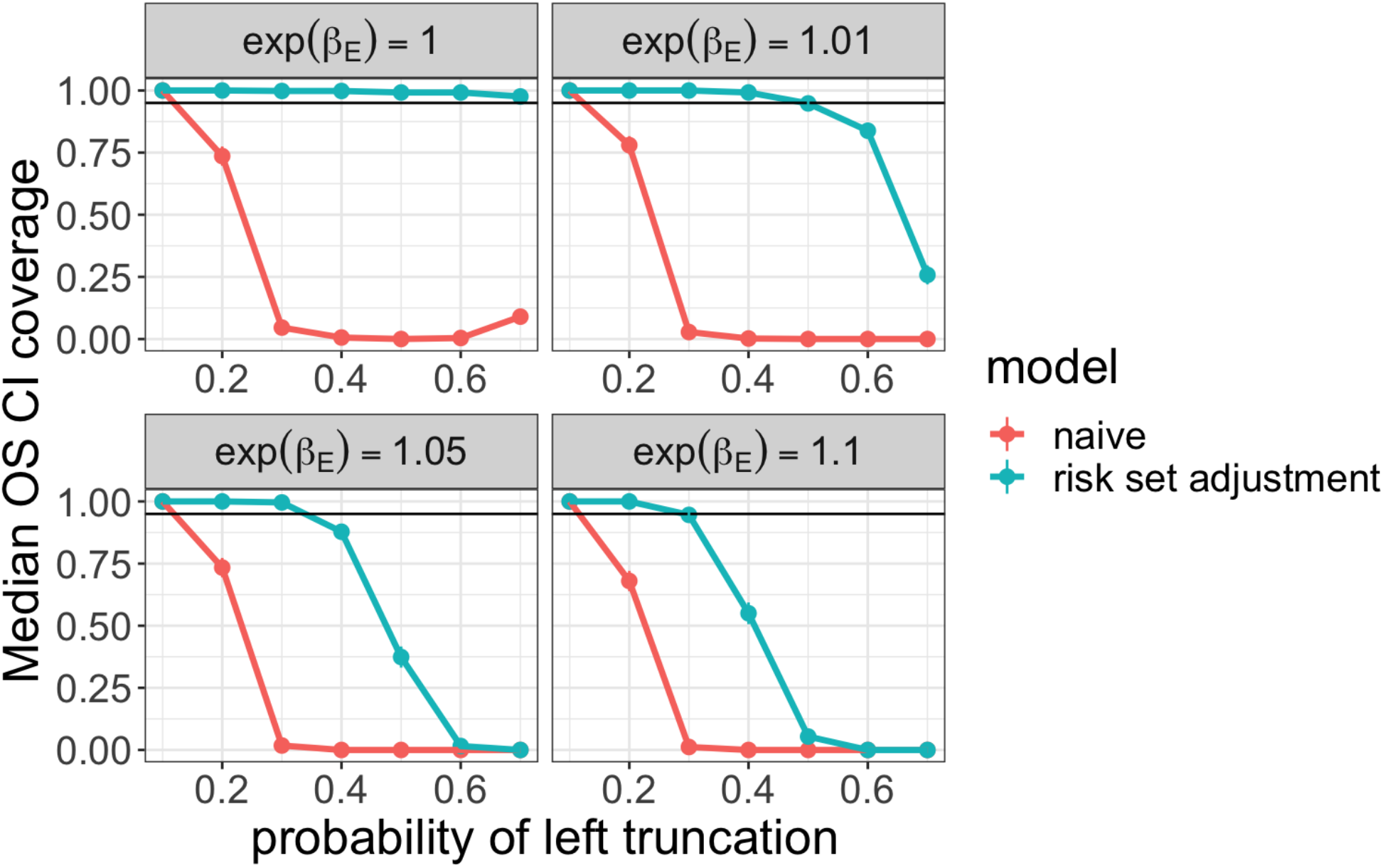
Coverage probability of 95% confidence intervals for median survival time in a single real world arm across simulation settings.

This is consistent with the relative bias results, which are displayed in Figure S4. We see that the bias is very high, given any moderate degree of truncation and any level of dependency.

**Figure S4.**
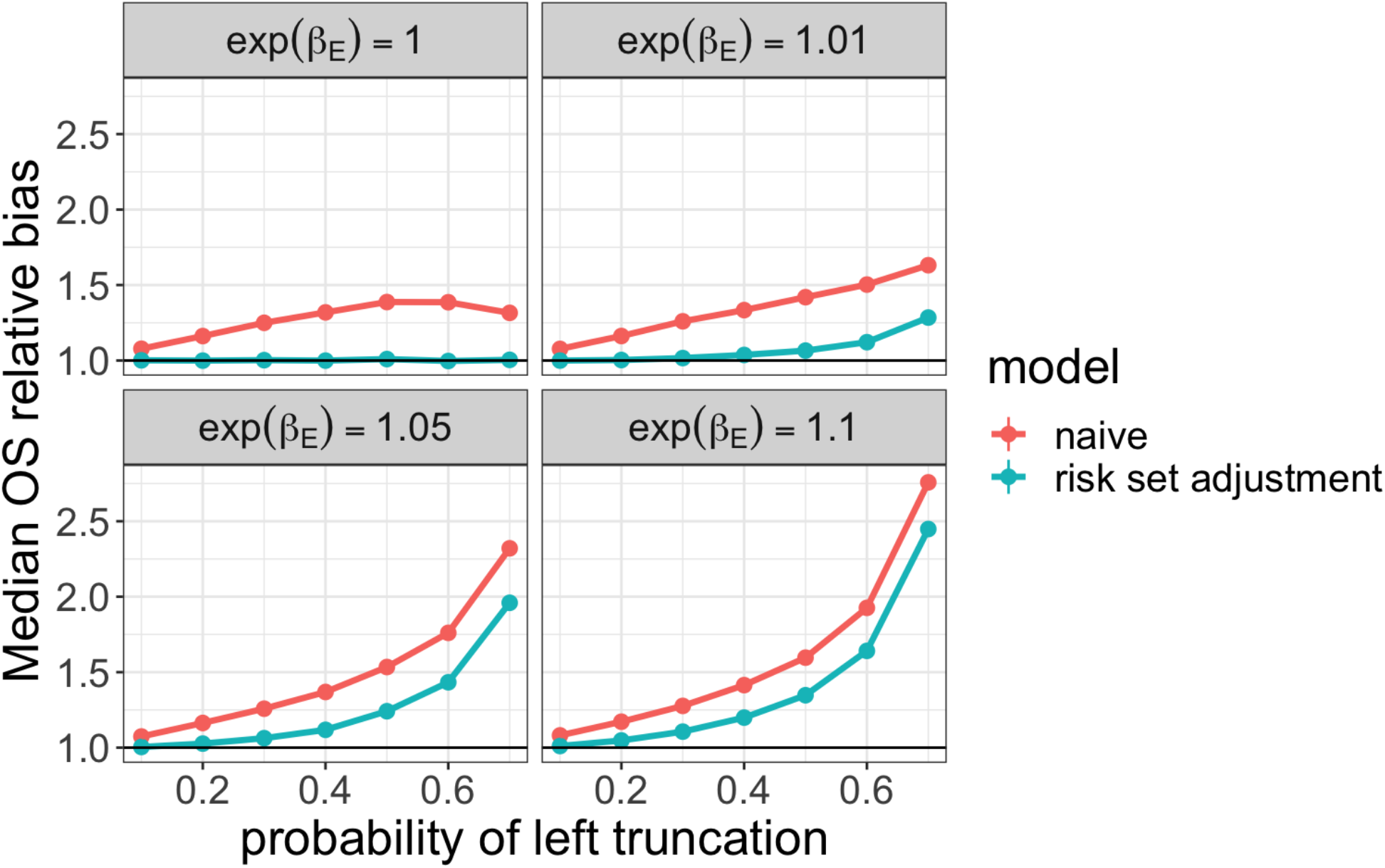
Relative bias for estimated median survival time in a single real world arm across simulation settings.

